# Blood immunomap for prediction of responses to aPD1 immunotherapy in metastatic non-small cell lung cancer

**DOI:** 10.1101/2024.08.27.24312529

**Authors:** Maria Semitekolou, Nikolaos Paschalidis, Domenico Lo Tartaro, Aikaterini Tsitsopoulou, Panagiota Stamou, Alexandros Mavroudis, Effrosyni Markaki, Athina Varveri, Ioannis Morianos, Matthieu Lavigne, Charalampos Fotsitzoudis, Sophia Magkouta, Konstantina Dede, Ioannis Kalomenidis, Konstantinos Samitas, Konstantinos Potaris, Andrea Cossarizza, Dimitrios Mavroudis, Sara De Biasi, Panayiotis Verginis

**Author notes:** Contributed equally (Maria Semitekolou, Nikolaos Paschalidis, Domenico Lo Tartaro). Correspondence: Panayotis Verginis, Nikolaos Paschalidis, Maria Semitekolou.

## Abstract

Immune checkpoint inhibitor immunotherapy has revolutionized the treatment of non-small cell lung cancer (NSCLC). Despite the immense success, still a significant proportion of patients do not develop durable responses, allowing disease progression accompanied by high mortality rates. Therefore, there is an imperative need for identification of reliable non-invasive predictive biomarkers to guide therapeutic decisions. Herein, we constructed a blood immunomap in NSCLC patients with metastatic disease, using a high-dimensional mass cytometry approach. Assessment of clinical responses to aPD1 immunotherapy revealed, among others, a significant expansion of CD8^+^PD-L1^+^ T cells in individuals not responding to immunotherapy. Of interest, CD8^+^PD-L1^+^ T cells were enriched in tumor biopsies and bronchoalveolar lavage of NSCLC individuals at early stages of disease as well as in pleural infusions of individuals with thoracic malignancies. Transcriptomic analysis revealed that CD8^+^PD-L1^+^ T cells exhibited a regulatory/exhausted phenotype, while various transcripts associated with the overall survival of NSCLC individuals, were mapped. Overall, our findings define an immunomap in the early stage and advanced NSCLC patients and identify immune-related events which may benefit the quest for identification of predictive biomarkers of immunotherapy responses.

## Introduction

Lung cancer represents the primary cause of cancer-related mortalities and morbidities worldwide with non-small cell lung carcinoma (NSCLC) accounting for 85% of lung cancer cases^1^. Regardless of the clinical accessibility of various treatments for NSCLC, including surgery, chemotherapy, radiotherapy, molecular targeted therapies, and immunotherapy, the 5-year survival rate remains unaffected due to the emergence of resistance to therapies. Genetic and epigenetic alterations, changes in the anti-tumor immune response pathways, tumor metabolic reprogramming, exhaustion of T-cell status and acquisition of a highly immunosuppressive tumor microenvironment (TME) are amongst the dominant mechanisms contributing to immunotherapy resistance^2,3^. Although the selection of the appropriate therapy for NSCLC patients largely remains stage dependent, the advent of immune checkpoint blockade (ICB) either as monotherapy or combination therapy, has reformed the management of locally advanced/metastatic NSCLC and has significantly improved our understanding regarding disease biology and mechanisms of tumor development and escape^4^. Even though ICB has demonstrated immense clinical success rate over other therapies, only a small proportion of NSCLC patients experience durable survival and overall clinical benefit^4^. The substantial number of clinical failures observed in combination treatments underlies the unmet need to delineate the mechanisms through which the tripartite of the host’s immune system, tumor and TME affects response or resistance to immune-based therapies and more importantly, design more rational ones^5^. Furthermore, the therapeutic benefit from currently available NSCLC treatments will be significantly intensified through the discovery and establishment of predictive biomarkers that will dictate patient’s response to ICB therapy^6^.

Despite the enormous expansion of therapeutic approaches in NSCLC, development of biomarkers that could guide personalized treatments, remain elusive. Current biomarkers, heavily rely on tumor biopsies and demonstrate low predictive value. To this end, intratumoral programmed death-ligand 1 (PD-L1) expression is considered a standard predictive biomarker for ICB efficacy and is the only one used in clinical routine^7–9^. Nevertheless, a conclusive association between PD-L1 expression and NSCLC patients’ overall survival has not been always witnessed^10^ and on top of that, an ICB treatment benefit was also observed in patients with < 1% PD-L1^11^. Besides, the discrepancies regarding the optimal conditions of PD-L1 immunohistochemistry assay, the cut-off value of staining positivity and the regional and temporal differences in PD-L1 expression question the predictive performance of PD-L1^7^. Tumor mutational burden (TMB) also emerged as an additional promising biomarker for predicting outcome to immunotherapy^12,13^. High TMB was associated with improved overall survival and extended PFS in advanced NSCLC patients independent of mismatch-repair deficiency, a mechanism that has been related to increased TMB^14–17^. Although initial reports were encouraging, latest studies argue the predictive value of TMB in NSCLC in regards to its arbitrary threshold^18^ and absence of positive correlation between TMB high phenotype and reliable survival advantage^19,20^. Considering that oncologists will have in the near future a large number of novel immunotherapies in the quiver, it is essential to identify robust prediction biomarkers that will improve treatment management in clinical practice. Although one could argue that the quest for predictive biomarkers should be focused on the tumor microenvironment, it is now established that this approach encompasses severe limitations. Among these the limited amount of tissue, the complications that occur during an invasive procedure, the under-representation of tumor heterogeneity and also the inaccessibility of tissue in difficult locations. Importantly, in majority of metastatic cases, in which tumor spread in various anatomic compartments, therapies are guided by the characteristics of the primary tumor which hold severe limitations. As blood is the most easily accessible tissue, the development of liquid biopsies has arisen as a promising, minimally invasive, and non-biased method to obtain sample for predictive biomarker identification^21^. Soluble and exosomal PD-L1, blood TMB, circulating non-coding RNA, peripheral blood (PB) cytokines, circulating free DNA and circulating immune cells are among numerous parameters being exploited as predictors of clinical benefit in NSCLC patients^22,23^. Despite the fact that numerous predictive circulating biomarkers have been evaluated in the last decades, very few, if any, met the demands of the clinic. Therefore, identification of circulating predictors of response to ICB therapy is urgently needed.

Herein, using a high-dimensionality mass cytometry approach, we created a blood immunomap in NSCLC patients with metastatic disease who underwent aPD1 treatment. Our findings identify increased frequencies of CD8^+^ PD-L1^+^ T cells in non-responders compared to responders to aPD1 immunotherapy. Of interest, as shown by Imaging Mass Cytometry (IMC) data, CD8^+^ PD-L1^+^ T cells were enriched in tumor biopsies, pleural infusions and BAL of NSCLC individuals, proposing this cells subset as candidate not only for the advanced but also for early disease detection. Transcriptomic analysis revealed that CD8^+^ PD-L1^+^ T cells exhibited an exhausted/anergic phenotype associated with their increased frequencies to non-responders to immunotherapy, and gene signatures correlated with the overall survival of an independent cohort. Overall, our findings reinforce the importance of circulating immune cells in the prediction of immunotherapy responses in NSCLC and propose that combination of liquid markers will substantially strengthen the discovery of robust biomarkers displaying superior clinical and predictive value.

## Results

### Study design and patient cohorts

A total of 56 PBMC samples from patients with unresectable, advanced (Stages III and IV) NSCLC were selected for CyTOF analysis (**Fig. 1a**, **Table 1** and **Supplementary Table 1**). The median age was 68 years with a male predominance (∼ 4.5 M:F ratio). Specific cell types were isolated with FACS, and transcriptomic analysis was performed with RNA seq (*n*=9). A second cohort of patients consisted of 19 NSCLC individuals (Stages I and II) from which heparinized peripheral blood and primary human lung tumor tissue was taken. All samples were analysed with CyTOF, and selected lung tumor tissue samples (n=2) were also analysed with Imaging Mass Cytometry (IMC). The median age was 69 years (∼ 1 M:F ratio). (**Fig. 1b**, **Table 2** and **Supplementary Table 2**). A third cohort of NSCLC patients (*n*=10) (Stages I-IV) was used for Bronchoalveolar lavage fluid (BALF) BALF analysis and a fourth one for Pleural Fluid (PF) (*n*=6). The median age was 65.4 years (∼ 1 M:F ratio) (**Fig. 1b Table 3, Supplementary Table 3** and **4**).

**Fig. 1.**
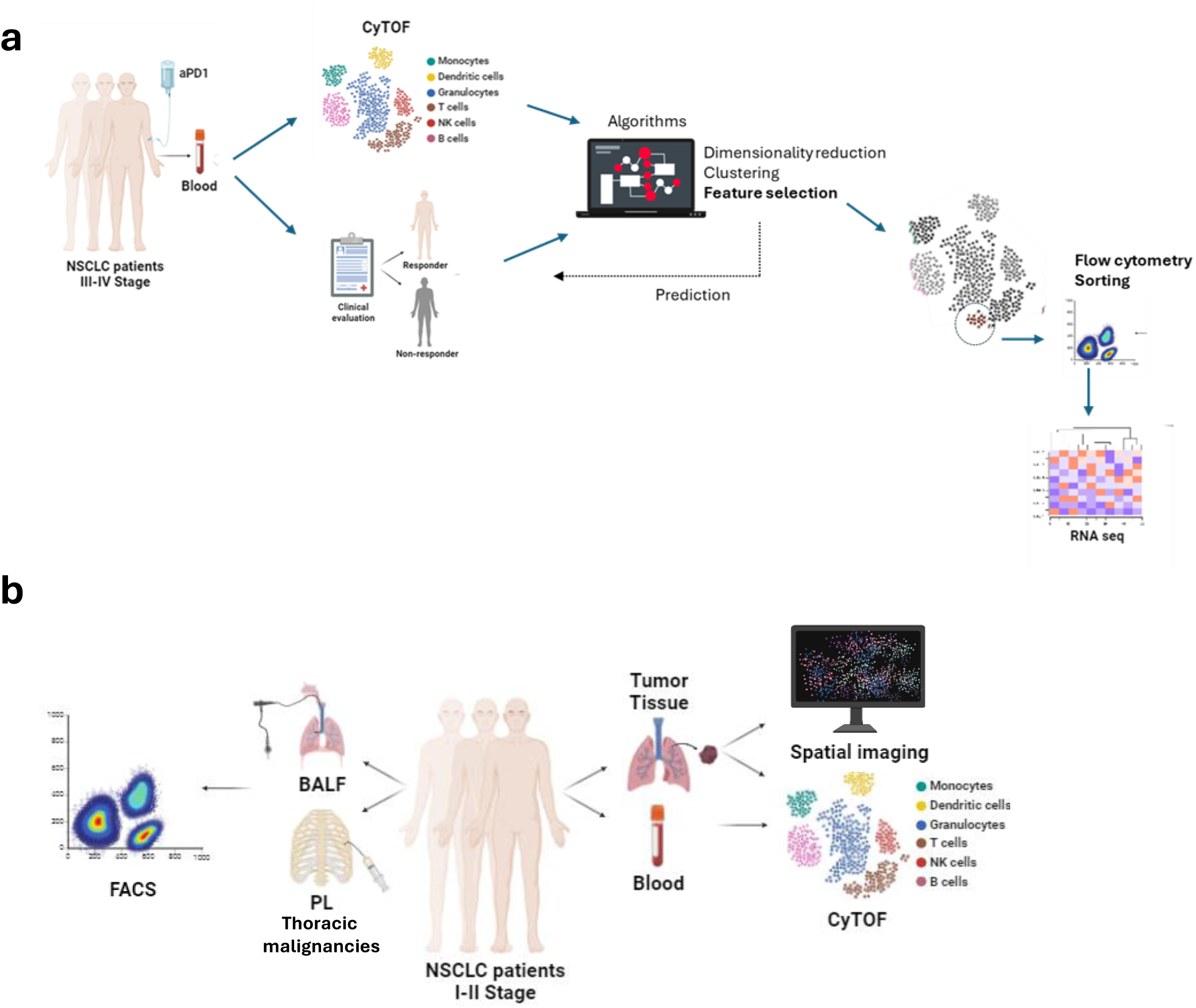
| Study outline. **a**) PBMCs from NSCLC patients (stage III-IV) before the initiation of aPD1 immunotherapy were analysed with CyTOF. Patients were clinically evaluated at 12 months and classified as responders (R) or non-responders (NR) to immunotherapy. Computational cytometry analysis tools were used to construct an immunomap and select key features for immunotherapy response prediction and further investigation with flow cytometry and RNAseq. **b**) PBMCs and lung tumor from NSCLC patients (stage I-II) were analysed with CyTOF and IMC. BALF and PF was also collected from individuals with thoracic malignancies, bronchiectasis or pleural infections for flow cytometry analysis. Created with BioRender.com.

### Single-cell Deep Immunophenotyping reveals a peripheral immunomap to distinguish advanced NSCLC individuals responding to aPD1 immunotherapy

Considering that blood constitutes the most accessible biological sample, we aimed to interrogate immune populations in the blood of patients with advanced NSCLC at baseline before the initiation of anti-PD1 immunotherapy. To this aim, we employed a high-dimensional, 33-marker, mass cytometry panel that allowed us to identify major immune cell populations such as T cells, B cells, NK cells and cells of the myeloid lineage as well as their subpopulations (**Supplementary Table 5)**. This analysis was based on FlowSOM, a clustering and visualization algorithm, based on a self-organizing map, employed to construct a peripheral blood immunomap to distinguish cell populations in an unsupervised way^24,25^.

Dimensional reduction and clustering analysis of PBMCs from these patients revealed a dominance of CD4 (T4) and CD8 (T8) T cells that were subdivided in distinct subsets reflecting activation and memory characteristics (**Fig. 2a-b**). We used the patterns of expression for markers CD45RA, CD45RO, CD127, CD25, CCR4 and CCR7 to annotate five clusters of T4 cells as *T4 N* (Naïve; CD45RA^+^, CD45RO^-^, CCR7^+^), *T4 TE* (Terminal Effector; CD45RA^+^, CD45RO^+^, CCR7^-^), *T4 CM* (Central Memory; CD45RA^-^, CD45RO^+^, CCR7^+^), *T4 EM* (Effector Memory; CD45RA^+^, CD45RO^-^, CCR7^-^) and TREG (T Regulatory cells; CD25^+^, CD127^low^, CCR4^+^). Similarly, we annotated four subsets of T8 cells as *T8 N*, *T8 TE, T8 CM and CD8 EM*. We also identified and annotated other major families of lymphocytes such as TCRγδ cells (*γδ-T*; CD3^+^, CD4^-^, CD8^low^, TCR *γδ* ^+^), double-negative T cells *(T DN*; CD3^+^, CD4^-^, CD8^-^, TCRgd^-^), NK-T cells *(NKT*; CD3^+^, CD4^-^, CD8^-^, TCR *γδ* ^-^, CD56^+^), B cells (*B N*; CD19^+^, CD20^+^, IgD^+^, memory B cells, *MBC* and activated B cell*, atBC*) and NK cells (*Early-NK*; CD56^+^CD16^-^ and *Late-NK*; CD56^+^CD16^+^). Furthermore, cells of the myeloid lineage clustered into seven subsets that were annotated as classical/intermediate monocytes (*Mono cl/int*, CD11c^+^, CD14^+^, CD16^-/lo^), *non-classical monocytes* (*Mono nc*, CD11c^+^, CD14^-^, CD16^+^), dendritic cell (*DC*; CD11c^-^, CD14^-^, HLA-DR^+^, IL-3R^+^), CD45 low expressing leukocytes (CD45^-/lo^ ^1^ ; CD66b^-^, CCR4^+^) and CD45^-/lo^ ^2^ (CD66b^+^, CD33^lo^, CD14^lo^). Patients were clinically evaluated based on RECIST 1.1 criteria and classified as responders (R) and non-responders (NR) 12 months post treatment initiation as shown in Table 1. When we compared the percentage of the identified immune cell signatures described above, we identified three clusters that were significantly different and in higher frequencies in non-responders (**Fig. 2c**). These were the *T8 CM* cells, the *CD45^-/lo^* ^2^ cluster and the pDC cluster.

**Fig. 2.**
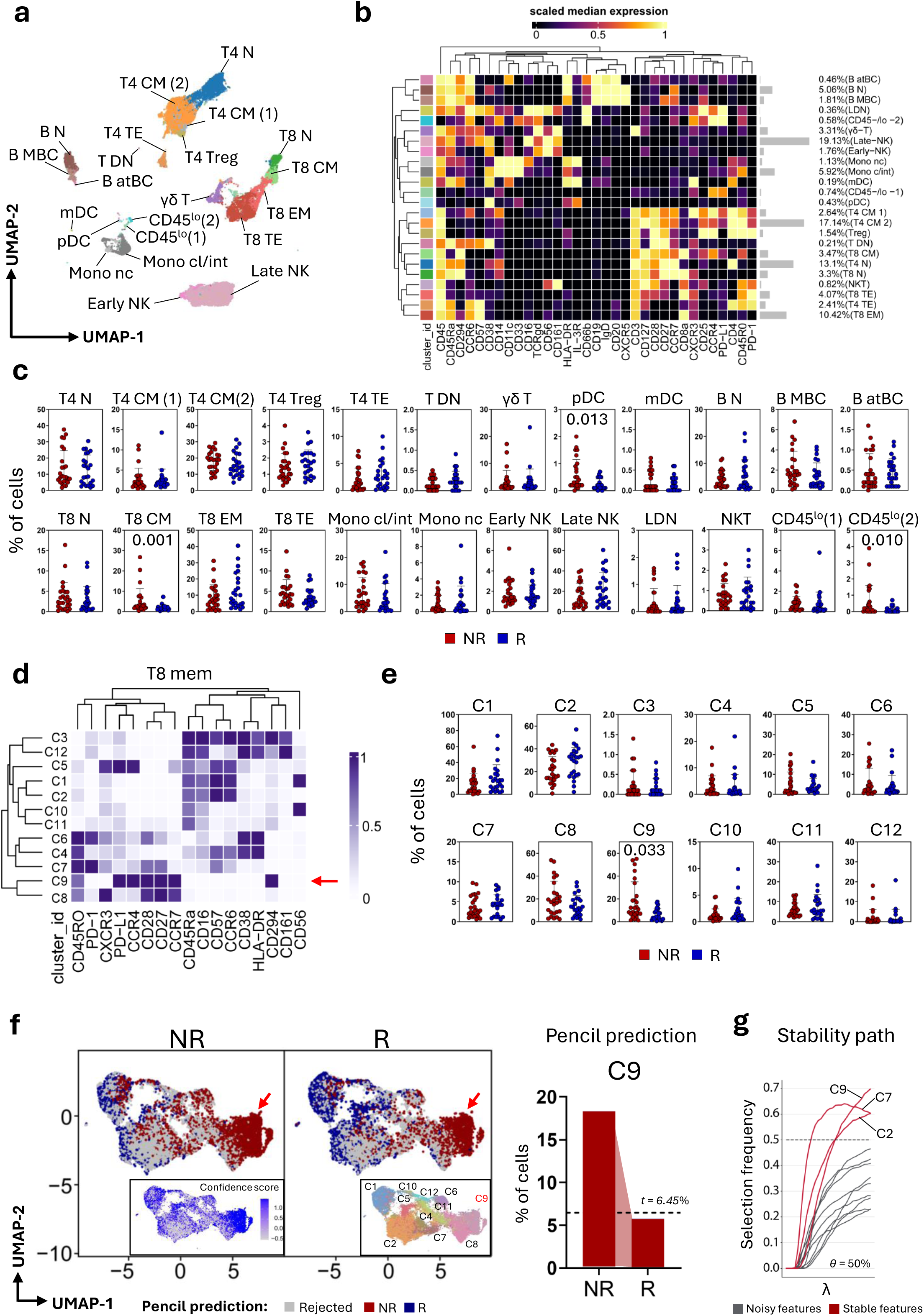
| Identification of peripheral blood immune signatures in NSCLC patients prior to the initiation of immunotherapy. **a)** UMAP plot of concatenated blood circulating CD45^+^ cells from responder (R; n = 25) and non-responder (NR; n = 25) samples from patients with NSCLC prior ICB. **b**) Heatmap of the median marker intensities of the 33 lineage markers across the 24 cell populations obtained using the FlowSOM algorithm after manual metacluster merging. The colors in the cluster_id column corresponds to those used for labeling the UMAP plot clusters as in **a**. The heatmap colors represent the median arcsinh marker expression (scaled 0–1) calculated from cells across all samples, blue indicating lower expression and red indicating higher expression. The light gray bar along the rows (clusters) and the values in brackets indicate the relative sizes of the clusters. **c**) Dot plots show the frequencies (% of live singlet PBMCs) of identified clusters among R (*n*=24) and NR (*n*=26). The central bar represents the mean ± SEM. Generalized linear mixed model (GLMM) test was used for the statistical analysis. Adjusted P-values are reported in the figure. **d**) Heatmap of the median marker intensities of the 17 lineage markers across the 12 cell populations obtained using the FlowSOM algorithm during the reclustering of T8 cells. **e)** Dot plots display the frequencies of T8 clusters in R and NR. The central bar represents the mean ± SEM. Statistical analysis was performed using a generalized linear mixed model (GLMM) test, with adjusted p-values reported in the figure. **f**) UMAP plot of R and NR T8 cells embedded with PENCIL prediction, confidence score of PENCIL prediction and FlowSOM clusters. In gray non classified cells (rejected), in blue cells associated with ICB response (R) and in red cells associated with ICB non-response. **g**) Stability path graphs denote clusters of T8 cells selected by StablL. The data-driven computed reliability threshold θ is indicated by a dotted line. Features selected by StablL (red lines) are shown

Since the most striking differences were observed within T8 cells, we decided to interrogate more in depth the phenotype of these cells by performing a new clustering step (see Methods). Clustering analysis revealed 12 clusters with distinct phenotypes (**Fig. 2d-e**). We found that the group of non-responders had significantly higher frequencies of T8 CM cells that expressed PD-L1 (Cluster *C9*, **Fig. 2e**).

Furthermore, to provide predictive insights, we employed a “phenotype-associated subpopulations from single-cell data (PENCIL), which is a supervised learning framework designed to enhance the accuracy and relevance of subpopulation analysis, based on rejection strategy learning^26^ ^27^. PENCIL prediction revealed that higher proportion C9, identified as T8 CM cells that expressed PD-L1 are enriched in blood of non-responder patients (**Fig. 2f**). To validate the prediction of PENCIL, we used Stabl, a general machine learning method that can identify a reliable set of biomarkers^28^. The Stabl model also identifies cluster C9 as the feature most associated with non-responsiveness to ICB therapy (**Fig. 2g**). These findings not only contribute to our understanding of the immune landscape in NSCLC, but also highlight specific cellular biomarkers that could predict patient outcomes following immunotherapy.

### Validation of the blood immunomap findings by flow cytometry

While several hematopoietic and non-hematopoietic cells express PD-L1 at various levels, cells of myeloid origin and tumor cells are considered as the main contributors of PD-L1 in cancer. Since our data suggest that PD-L1-expressing CD8^+^ T cells could predict responses to aPD1 immunotherapy we sought to validate these results through flow cytometry analysis. To this end, gating on CD3^+^ T cells no major differences were observed in frequencies of total CD4^+^ and CD8^+^ T cells between the two groups of NSCLC individuals. Notably, assessment of PD-L1 expression, revealed a significant increase of PD-L1-expressing CD8^+^ T cells in blood of non-responders compared to responders (**Supplementary Fig. 1a**) confirming the mass cytometry results, whereas no significant differences in frequencies of PD1-expressing CD8^+^ T cells were observed between the two groups (**Supplementary Fig. 1b**). We also analysed monocytic cell subsets that abundantly express PD-L1 and although no significant alterations in frequencies of CD16^-^ CD14^+^ classical monocytes (CL), CD16^+^ CD14^+^ intermediate monocytes (INT) and CD16^+^ CD14^-^ non-classical monocytes (NCL) were observed between responders and non-responders (**Supplementary Fig. 1c-e**), PDL-1-expressing NCL cells were significantly increased in non-responders while no difference was observed in regard to PD-L1-expressing CL population. To conclude, these results validate the blood immunomap findings in NSCLC individuals and proposes that PD-L1-expressing CD8^+^ T cells may contribute to the prediction of response to aPD1 immunotherapy.

### PD-L1-expressing CD8+ T cells are enriched in the tumor biopsies from NSCLC individuals

Following the identification of circulating PD-L1-expressing CD8^+^ T cells as a potential predictive biomarker to aPD1 immunotherapy in advanced NSCLC individuals and considering that immune checkpoint immunotherapy obtained approval as first and second line therapies, we asked whether PD-L1^+^CD8^+^ T cells could be identified among tumor infiltrating leucocytes (TILs) at early stages of tumor development. For this, TILs and blood were collected form NSCLC patients (**Table 2 and Supplementary Table 2**) subjected to mass cytometry analysis using the same 33-marker panel as above. We used a similar analysis approach, like the one shown on the peripheral blood immunomap, that included cluster identification with FlowSOM, manual annotation based on canonical markers as well as markers indicative of cell activation and differentiation state and projection of the results on dimensionality reduction maps (**Fig. 3a-b**).

**Fig. 3.**
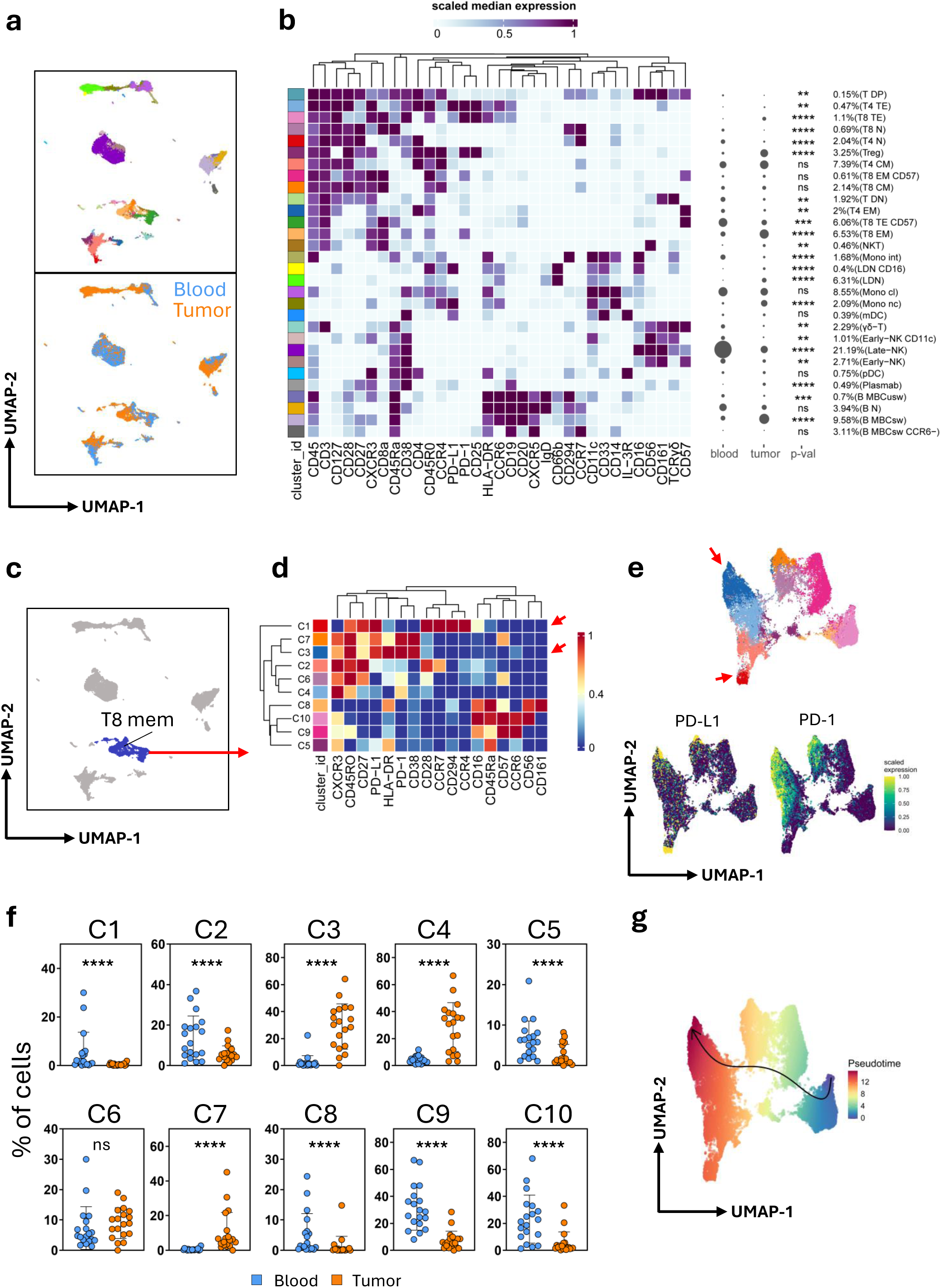
| Distinct subsets of CD8^+^ PD-L1^+^ cells are present in the periphery and lung tissue of NSCLC patients. **a)** UMAP plot of concatenated CD45^+^ T cells from peripheral blood (n = 38) and tumor (n = 38) samples from early-stage NSCLC patients color-coded by FlowSOM clusters (Top panel) or tissue origin (Bottom panel). **b)** Heatmap of the median marker intensities of the 33 lineage markers across the 32 cell populations obtained using the FlowSOM algorithm after manual metacluster merging. The colors in the cluster_id column correspond to those used for labeling the UMAP plot clusters as in **a**. The heatmap colors represent the median arcsinh marker expression (scaled 0–1) calculated from cells across all samples, with white indicating lower expression and dark purple indicating higher expression. The light gray balloon plot along the rows (clusters) indicates the relative sizes of the clusters among blood and tumor tissue. **c**) UMAP plot as in **a** depicting the selected T8 memory cells (blue dots). **d**) Heatmap of the median marker intensities of the 17 lineage markers across the 10 cell populations obtained using the FlowSOM algorithm during the reclustering of T8 memory cells. **e**) UMAP plot of T8 memory cells, with the top panel color-coded by FlowSOM clusters and the bottom panel color-coded by PD-1/PD-L1 expression. Red arrows indicate the C1 and C3 clusters expressing PD-L1. **f**) Dot plots display the frequencies of T8 memory clusters in peripheral blood and tumor. The central bar represents the mean ± SEM. Statistical analysis was performed using a generalized linear mixed model (GLMM) test, with adjusted p-values reported in the figure as *p <0.05, **p <0.01, ***p<0.005, ****p<0.001. **g**) UMAP plot as in **e** depicting the trajectory analysis (black arrow) of T8 memory cells color-coded by pseudotime value.

Our comparative analysis between peripheral blood and TILs from NSCLC patients revealed distinct immune landscape, reflective of the complex interplay within the tumor microenvironment.

We observed that lung TILs were dominated by effector T8 cells, T4 cells, as well activated B cells (clusters *T8 TE*, *T8 EM*, T4 TE, *B EFF*, *B MBscw* and *Plasmab*) as opposed to the peripheral blood where naïve lymphocytic populations mostly prevailed (clusters *T4 N*, *T8 N* and *B N*) (**Fig. 3b**). In the TILs, the T4 compartment was characterized by an elevated relative frequency of regulatory T cells (*TREG*), suggesting a pronounced immunosuppressive environment that may impede effective anti-tumor responses. We observed no significant difference in the proportions of central memory (*T4 CM*) subsets within this compartment, pointing to a selective enrichment of Tregs. Moving to the T8 compartment, TILs exhibited a marked increase in terminal effector cell (*T8 TE*) frequencies, which could indicate a heightened, albeit potentially thwarted, cytotoxic response within the tumor milieu. We also observed marked differences in other immune cell populations, including NK cells, (*Early-NK* and *Late-NK*), as well as most monocyte subsets. Moreover, non-classical monocytes (*Mono nc*) were more represented within the TILs, potentially signifying a unique migratory or survival pattern in the tumor context. (**Fig. 3b**).

Focusing on CD8^+^ T cells across peripheral and tumor environments, we observed an emerging dichotomy in the differentiation stages and expression patterns of PD-1 and PD-L1 (**Fig. 3c-f**). Among PBMC, we observed higher percentages of central memory cells expressing PD-L1 and CD294 (C1; CD45RA^-^, CD45RO^+^, CCR7^+^, PD-L1^+^, CD294^+^), as well as terminal effector cells with variable CD57 expression (C8, C9, C10; CD45RA^+^, CCR7^−^), which exhibit notably low levels of PD-L1. This profile contrasts sharply with the T8 compartment in TILs, which is predominantly composed of effector memory subsets (C3, C4, and C7; CD45RA^−^, CD45RO^−^, CCR7^−^). Moreover, C3 and C7 clusters were highly positive for both PD-1 and PD-L1 (Fig. 3d-f). Pseudotime trajectory analysis confirmed that C3 and, to a lesser extent, C1 are at the terminal stage of the differentiation pathway, likely indicating their exhausted status (**Fig. 3g**).

### Tumor infiltrating PD-L1+ CD8+ T cells interact with myeloid and cancer cells and exhibited exhausted characteristics

Given that CD8^+^ PD-L1^+^ T cells have been found in both blood and the TME, we aimed to determine their localization within NSCLC. To analyze the cellular composition of NSCLC tumors while preserving spatial context, we employed imaging mass cytometry (IMC) to detect 22 proteins in two formalin-fixed, paraffin-embedded samples (**Supplementary Table 5)**. We identified 124,765 cells in total, and using an unsupervised lineage assignment approach, we classified these into 15 distinct cell populations (**Fig. 4a-b**). Four of these populations (C6, C8, C12, C15) expressed Pan-keratin (Pan-K) along with E-cadherin (E-Cadh) and were classified as tumor cells (**Fig. 4b**). Cells expressing alpha smooth muscle actin (α-SMA; grouped into C2) were classified as stromal cells (**Fig. 4b**). Seven clusters of myeloid cells have been identified: dendritic cells (C11) expressing CD11c, macrophages (C1, C3, C5, C9, C13) expressing CD68, and monocytes (C14) expressing HLA-DR and intermediate levels of CD16 (**Fig. 4b**). Within the lymphoid compartment, we observed a cluster of B-T cells (C4) forming ectopic structures known as tertiary lymphoid structures (TLS) and a cluster of Foxp3 positive Treg cells (C10) (**Fig. 4b**). Additionally, we identified a cluster of CD8^+^ T cells expressing intermediate levels of PD-1 and PD-L1 (C7). Upon manual assessment of the IMC data, we confirmed the presence of PD-L1^+^ CD8^+^ T cells within the lung cancer tissue. Notably, these cells were found in close proximity to cancer cells (Pan-K^+^; green) or within TLS (CD20^+^, cyan) (**Fig. 4c**). To specifically characterize the interactions and communication patterns of PD-L1^+^ CD8^+^ T cells with other cell clusters, we quantified cell–cell spatial relationships. We reported strong interactions between PD-L1^+^ CD8^+^ T cells (C7) and macrophages/monocytes (C3, C9, and C14), stromal cells (C2), cancer cells (C6 and C8), and to a lesser extent with B-T cells (C4) (**Fig. 4d**).

**Fig. 4.**
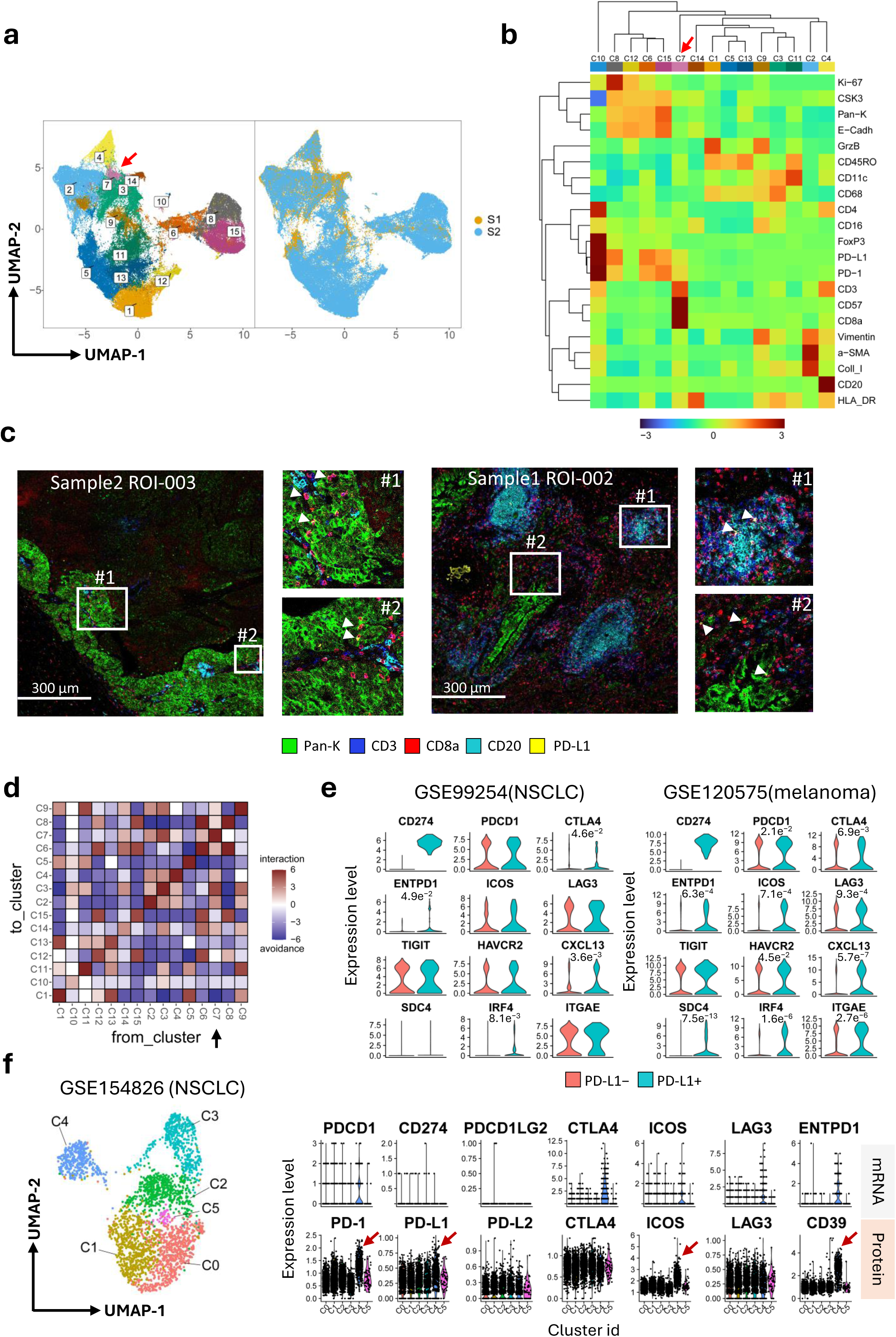
| Single-cell spatial localization of lung infiltrating PD-L1^+^ CD8^+^ T cells. **a)** Two-dimensional UMAP representation of multiplexed proteomic data highlighted by cell phenotype (clusters; n =15) and sample id (n=2; total ROI = 6). Each dot represents one cell. **b)** Heatmap of median values of normalized protein expression per cell cluster. Markers and clusters were arranged by hierarchical clustering with Ward’s method. **c)** Representative multichannel IMC images (ROI: region of interest) from sample 2 (left) and sample 1 (right). Pan Keratin (Pan-K, green), CD3 (blue), CD8a (red), CD20 (cyan) and PD-L1 (yellow) were used to depict the structure of the tumor tissue along with the localization of TLS and PD-L1^+^ CD8^+^ T cells. **d)** Heatmap depicting significant pairwise cell–cell interaction (red) or avoidance (blue) between all cell types of the dataset. **e**) Violin plot showing the levels of manually selected exhaustion genes in PD-L1^−^ vs. PD-L1^+^ CD8^+^ T cells from NSCLC (GSE99254, stage I-IV) and melanoma (GSE120575, stage III-IV) cohorts. **f**) (left) UMAP depicting CD8^+^ T-cell heterogeneity in the GSE154826 dataset (NSCLC patients, stage I-IIB). Cells are colored according to the six clusters defined in an unsupervised manner; (right) violin plot illustrating both mRNA and protein expression of manually selected markers indicative of T cell dysfunction.

**Fig. 5.**
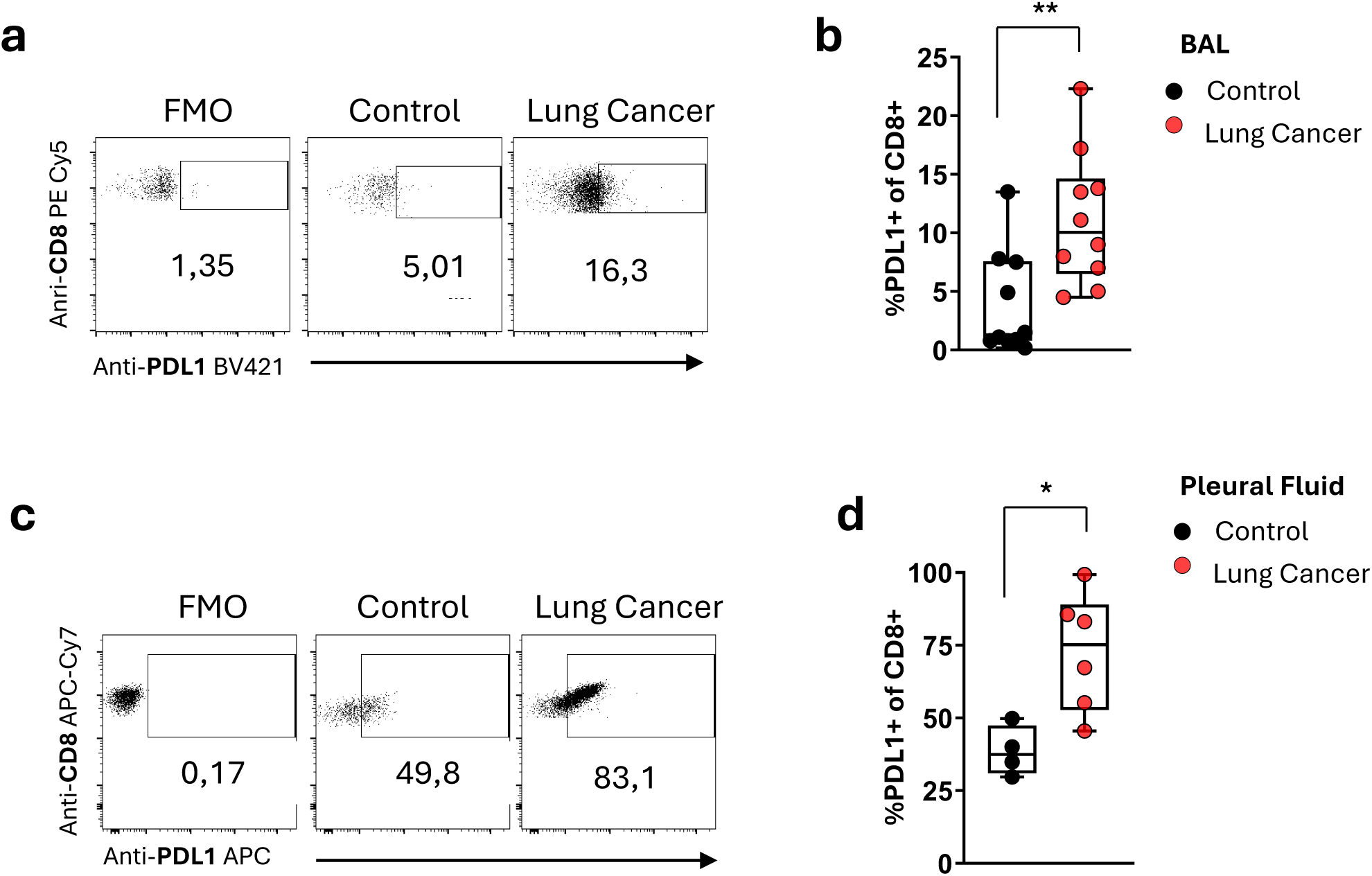
| Flow cytometry analysis reveals increased percentages of PD-L1^+^ CD8^+^ T cells in the bronchoalveolar lavage (BAL) of NSCLC individuals and in pleural fluid (PF) of individuals with thoracic malignancies. **a)** Representative FACS Dot plots showing gating of CD8^+^ PD-L1^+^ T cells in BALF from NSCLC individuals with bronchiectasis (n=10) **b)** Box and whiskers plots of collective flow cytometry data of CD8^+^ PD-L1^+^ cells in BALF. **c)** Representative FACS Dot plots showing gating of CD8^+^ PD-L1^+^ T cells in PF from individuals with thoracic malignancies (n=6) and pleural infections (n=4) bronchiectasis (n=10 per group) **d)** Box and whiskers plots of collective flow cytometry data of CD8^+^ PD-L1^+^ cells in PF cells. Unpaired two tailed Mann-Whitney test *p <0.05, **p <0.01.

To validate the presence of PD-L1^+^ CD8^+^ T cells in both NSCLC and various other solid tumors, we performed additional investigations using three publicly accessible single-cell RNA sequencing (scRNA-seq) datasets (see Methods). In the first two datasets, which included cells from NSCLC and melanoma patients, we identified CD8^+^ T cells expressing CD274, the gene encoding *PD-L1*. These cells exhibited higher expression levels of genes associated with an exhausted phenotype, such as *CTLA4, PDCD1 (PD-1), ICOS, LAG3, ENTPD1 (CD39), HAVCR2 (TIM-3), SDC4, IRF4 and CXCL13*, compared to PD-L1 negative CD8+ T cells (**Fig. 4e**). We also considered a third dataset comprising cells from patients with resectable NSCLC, analyzed using CITE-seq technology to integrate protein and transcriptome measurements at a single cell level. CD8^+^ T cells were selected and analyzed using an unsupervised graph-based clustering approach, revealing six subpopulations (**Fig. 4f**, **left**). Among these, cluster 4 (C4) was the only one expressing exhausted genes such as *PDCD1, CTLA4, LAG3, and ENTPD1*. Due to the low sequencing depth of CITE-seq data (∼0.1 million paired-end reads per cell for CITE-seq compared to a median of ∼1.4 million paired-end reads per cell for Smart-seq2), we were unable to detect PD-L1 at the transcript level (**Fig. 4f** right). However, at the protein level, we observed higher expression of PD-L1 in cluster 4, along with PD-1, ICOS, and CD39. These data suggest that PD-L1^+^ CD8^+^ T cells appear more exhausted compared to their PD-L1^−^ counterparts (**Fig. 4f** right).

### Identification of PD-L1^+^CD8^+^ T cells in tissues associated with NSCLC diagnosis

Since there is a significant gap in our knowledge on early diagnosis of NSCLC, together with the immunomap results which identify enriched frequencies of PD-L1^+^ CD8^+^ T cells in lung tumor tissues, we asked whether such cells could also be detected in bronchoalveolar lavage (BAL) and pleural fluid (PF) tissues that are easily accessible, semi-invasive and routinely monitored for diagnosis of lung cancer. To this end, using flow cytometry, we observed increased frequencies of PD-L1^+^ CD8^+^ T cells in the BAL of NSCLC individuals who underwent bronchoscopy compared to individuals with bronchiectasis serving as disease control (**Fig.5a-b**). In line with this, increased frequencies were also detected in PF of individuals with thoracic malignancies in comparison with individuals with pleural infection who served as the respective control group (**Fig.5c-d**). These findings raise the possibility of PD-L1^+^ CD8^+^ T cells to be considered in the diagnosis of NSCLC.

### Peripheral PD-L1^+^ CD8 T cells from non-responder patients represent a markedly dysfunctional subset

To shed light on the molecular identity of PD-L1^+^ CD8^+^ T cells, we performed transcriptomic analysis on sorted cells (**Supplementary Fig. 2**) and compared their gene signatures to those of PD-L1^−^ CD8^+^ T cells obtained from the PBMCs of NSCLC individuals(see Methods In total, we identified 317 DEGs (FDR < 0.05). PD-L1^+^ CD8^+^ T cells expressed higher levels of genes associated with T cell immune suppression or exhaustion, including *NR4A1, IGFBP2 and LAYN*^29–31^, (**Fig. 6a**). In contrast, PD-L1^−^ CD8^+^ T cells seems to be largely distinct, featuring elevate expression of genes promoting CD8^+^ stemness such as *WNT1*^32^, effector functions such as *SPRY1*^33^ and ATP generation such as *PFKFB1* or *ATP8B4*^34^ (**Fig. 6a**).

**Fig. 6.**
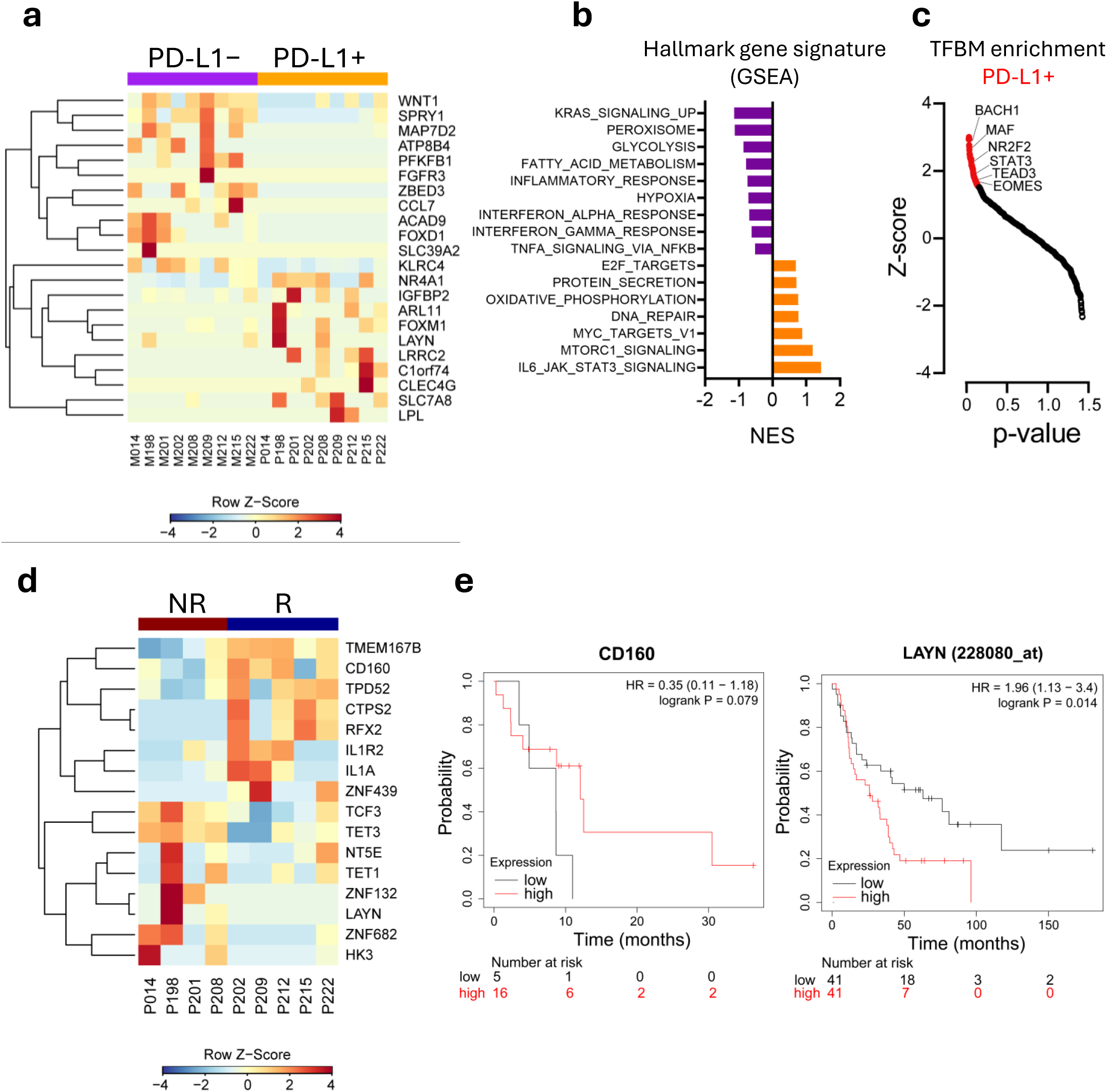
| Transcriptional profiling of PD-L1^+^ CD8^+^ T cells. **a)** Heatmap of differentially expressed genes (FDR < 0.05) between FACS-sorted PD-L1^−^ (n = 9) vs. PD-L1^+^ (n = 9) CD8+ T cells subsets from the blood of patients with NSCLC, as obtained by bulk RNA-seq. Selected differentially expressed genes are indicated. **b)** Hallmark gene sets (MsigDB; as obtained by GSEA) significantly enriched in cells sorted as in **a**. **c)** Transcription factor binding motif (TFBM) enrichment analysis by pScan of RNA-seq data obtained as in (**a)**. Only genes upregulated in PD-L1^+^ CD8^+^ T cells were used. Colored dots indicate significant hits. **d)** Heatmap of differentially expressed genes (FDR < 0.05) between FACS-sorted PD-L1^+^ CD8^+^ T cells from responder (n = 5) and non-responder (n = 4) NSCLC patients receiving ICB therapy. Selected differentially expressed genes are indicated. **e)** Kaplan–Meier overall survival (OS) curves in the immunotherapy dataset (KM plotter) of patients with NSCLC. The mean z-score value of reported genes was used to classify tumor samples into LOW and HIGH expression groups. P-values were calculated using the log-rank (Mantel–Cox) test. Hazard ratio (HR) and log-rank p-values are reported in the figures.

Gene set enrichment analysis (GSEA) further revealed that PD-L1^−^ CD8^+^ T cells were characterized by transcripts associated with glycolysis, fatty acid metabolism, inflammatory response and interferon signaling pathways, whereas PD-L1^+^ CD8^+^ T cells were characterized by the expression of genes associated with the oxidative phosphorylation signaling pathway, DNA repair and other mechanistic correlates of dysfunction, including MYC targets (**Fig. 6b**). To identify transcriptional regulators of PD-L1^+^ CD8^+^ T cells, we performed computational analysis of transcription factors (TF)-binding motif enrichment at the promoters (–950, +50 bp from the TSS) of DEGs obtained from bulk RNA-seq data. This analysis showed enrichment of binding motifs of TFs, including *MAF* and *EOMES*, both promoting CD8 T cell exhaustion^35,36^,. We additionally identified an enrichment for *STAT3* and *TEAD3*, involved in inhibition of interferon expression, and induction of PD-L1 expression^37^ (**Fig. 6c**).

Given that higher percentages of PD-L1^+^ CD8^+^ T cells have been found in peripheral blood (PB) of non-responder patients, we compared the transcriptomes of these cells to those from responders to determine if they were transcriptionally different (see methods). Overall, we identified 85 differentially expressed genes (supplementary table2). Cells from responders expressed high amounts of transcripts encoding molecules involved in effector function such as *CD160*, while cells from non-responders were characterized by exhausted and immunosuppressive genes such as *LAYN*, *TET1* and *TET3*^30,38^ (**Fig. 6d**). We then evaluated whether these genes possess prognostic value and correlate with better overall survival (OS) in NSCLC patients undergoing ICB therapy. Our analysis revealed that higher levels of *CD160* expression were significantly associated with improved OS (P = 0.079), whereas elevated levels of *LAYN* were linked to poorer OS (P = 0.014; **Fig. 6e**).

## Discussion

Despite enormous efforts over the last decade to identify tissue or blood biomarkers to guide prognosis to ICI therapy in the treatment of NSCLC individuals, several limitations have hampered the clinical application of the suggested panels, leaving an important gap in the prediction of ICI responses. Herein, focusing on blood of NSCLC individuals with advanced disease, we structured an immunomap through a high-dimensional CyTOF approach which assisted to identify immune cell subsets with different representation in responders versus non-responders NSCLC individuals to aPD1 treatment. Our findings identify increased frequencies of CD8^+^ PD-L1^+^ T cells in periphery of non-responders compared to individuals that respond to ICI therapy. Notably, CD8^+^ PD-L1^+^ T cells were enriched in tumor biopsies, pleural infusions, and BAL of NSCLC, placing this cell subset as a candidate for the early detection of NSCLC.

Expression of PD-L1 by T cells has been described in various cancer settings. To this end, Diskin et al described increased frequencies of CD4^+^ and CD8^+^ T cells expressing PD-L1 in a mouse model of pancreatic ductal adenocarcinoma (PDA), while functional assessment of PD-L1 expression was performed only using CD4^+^ T cells ^39^ . In human settings, using an ex vivo lymph node assay, circulating PD-L1-expressing T cells (both CD4^+^ and CD8^+^) were shown to be prognostic on overall and progression-free survival in unresectable stage III and IV melanoma cohort^40^, while in the same study CD8^+^ PD-L1^+^ T cells were proposed as a marker of resistance to aCTLA-4 treatment. In line with this, CD8^+^ PD-L1^+^ T cells were significantly increased in blood of melanoma patients close to experience disease relapse or disease-related death^41^. In lung cancer, levels of PD-L1 expression were evaluated on circulating CD3^+^, CD4^+^ and CD8^+^ T cell subsets, and was shown that NSCLC patients possessed increased frequencies of CD8^+^ PD-L1^+^ T cells compared to healthy individuals, and survival was decreased among patients with high frequencies of those cells^42^. Furthermore, PD-L1^+^ CD8^+^ T cells were enriched in primary lesion of a NSCLC cohort and exerted an immunosuppressive function of effector cytotoxic T cells^43^. However, whether such cells could predict immunotherapy responses in NSCLC has not been addressed. Our extensive phenotypic characterization revealed that circulating CD8^+^ PD-L1^+^ T cells constitute a heterogeneous population and the subset of cells which correlate with aPD1 resistance and disease progression expresses markers such as CD127 and CD28, intermediate levels of CD25 while did not express PD-1, HLA-DR, CD38 and CD57.

Attempting to shed light on the functional importance of these cells, we first focused on the phenotypic markers. Thus, the levels of CD127 expression are determined by its ligand IL-7, in which absence of IL-7 signalling results in increased CD127 expression by CD8^+^ T cells and associated with attenuated cytotoxic potential and exhausted phenotype in melanoma patients^44^. Expression of CD38 by intratumoral CD8^+^ PD-L1^+^ T cells, was shown to correlate with poor responses to aPD1 immunotherapy in preclinical models and melanoma patients^45^ suggesting an immunosuppressive role of this subset in the TME. Accordingly, we identify CD38^+^PD-L1^+^CD8^+^ TILs which are diminished in the periphery of the same NSCLC cohort. In contrast, the subset of CD8^+^ PD-L1^+^ T cells which is enriched in the periphery of non-responders to aPD1 ICI individuals, lack CD38 and HLADR. It was demonstrated that CD38^+^ and HLADR^+^ CD8^+^ T cells exhibit cytotoxic activity and inflammatory cytokine production in patients with infectious diseases and they decline upon resolution of the infection suggesting an effector T cell subset^46^. Although this literature allows the formulation of the hypothesis that peripheral CD38^-^ HLA-DR^-^ CD8^+^PD-L1^+^ T cells lack an effector function and may be anergic/exhausted/regulatory, direct evidence is still missing. Surprisingly, in support to this hypothesis, the peripheral CD8^+^ PD-L1^+^ T cells in non-responders in our study do not express PD1, which although in infectious diseases is considered as a surrogate marker of exhaustion, in tumor settings PD1 expression characterizes the activation stage of CD8^+^ T cells^47,48^ suggesting that lack of PD1 expression may indicate a dysfunctional T cell subset. Finally, the lack of CD57 expression by CD8^+^ PD-L1^+^ T cells points towards an immunosuppressive stage of this subsets if we consider that CD57 expression correlated with expression of perforin and granzymes by CD8^+^ T cells which are essential for their cytotoxic activity^49^ . Overall and based on this extensive phenotype it is difficult to ascribe a functional state on CD8^+^PD-L1^+^ cells, it is possible that they exhibit a dysfunctional state similar to exhausted or regulatory T cells. This is further supported by the transcriptomic analysis of these cells, which revealed genes associated with T cell dysfunction, such as *NR4A1, IGFBP2 and LAYN*^29–31^, to be upregulated in CD8^+^PDL1^+^ compared to CD8^+^PDL1^-^ cells. Mechanistically, CD8^+^PDL1^+^ T cells, may directly suppress CD4^+^ or CD8^+^ T cells responses upon engagement of PD-L1 to PD1 expressed by effector cells^39^ or other inhibitory receptors such as the inhibitory killer cell immunoglobulin-like receptors (KIRs)^50^, may inhibit antigen presentation considering that myeloid cells express PD1^51^, and also could downregulate PD1 expression to avoid targeting of aPD1 ICI immunotherapy and to promote tumor immune evasion. The latter is further supported by our findings which reveal that CD8^+^PDL1^+^ T cells strongly interacted with monocytes in the TME. In addition to monocytes, our imaging cytometry data reveal a favourable interaction of CD8^+^PDL1^+^ T cells with cancer cells and stromal cells. How those interactions imprint on the functional status of CD8+ T cells expressing PDL1 remains unknown. Nevertheless, stromal cells recently showed to promote a tumor-permissive TME through formation of synapsis with Treg cells^52^ and thus interaction with CD8 T cell subsets could also envisioned.

Using the same immunomap approach in an independent cohort of NSCLC patients at the early stages of disease, several CD8^+^ T cells subsets were identified to express PD-L1 among TILs. CD8^+^ PD-L1^+^ TILs cells with a similar phenotype to the circulating counterparts, observed in metastatic NSCLC patients were in lower frequencies but still significantly higher at the periphery of this independent cohort. CD8^+^ TIL subsets express even higher levels of PD-L1, but express high levels of CD38 and HLA-DR while they do not express CD57. Nevertheless, considering the plasticity of the T cell compartment, including the CD8^+^ T cells, whether these phenotypes reflect “permanent” functional cell states and if they adopt a different phenotype upon exposure to TME remail to be determined.

In spite of the urgent demand for the identification of non-invasive predictive biomarkers for ICI immunotherapy responses and the enormous number of studies which propose potential blood biomarkers, their translation into clinical practice remains an unmet need. Most likely the “perfect” biomarker will consist of a panel of molecules and genes expressed by hematopoietic and non-hematopoietic cells if we consider that such cells circulate and eventually populate the TME. Our efforts towards this direction, focused on the construction of a blood immunomap of advanced NSCLC individuals responding or not to aPD1 ICI immunotherapy. We identified CD8^+^ PD-L1^+^ T cells to dominate in non-responder cells while these cells were enriched among TILs, in pleural infusions and BAL of independent disease cohorts. Incorporation of our findings to existing knowledge together with studies in larger cohorts and independent validations may pave the way for development of predictive and/or prognostic non-invasive biomarkers for prediction of ICI responses in NSCLC individuals.

## Methods

### Patient samples

This study was based primarily on three cohorts of NSCLC individuals, one cohort of individuals with malignant pleural effusions, one with pleural infection and one with bronchiectasis. The first cohort entailed individuals with unresectable, advanced (Stages III and IV) NSCLC (*n*=56), recruited through the Department of Medical Oncology at the University General Hospital of Heraklion. Heparinized peripheral blood was obtained from this cohort and donors were treatment naïve at the time of surgery. Of the 56 individuals with advanced NSCLC, 36 were histopathologically annotated as lung adenocarcinoma, 17 as squamous cell carcinoma, 2 as pleomorphic carcinoma and 1 as adenocarcinoma and squamous cell carcinoma. Clinical characteristics including age, sex, smoking status, pathological subtypes and stages, oncogenic driver mutations, PD-L1 expression, line of therapy, treatment and mortality on all donors are listed in Table 1 and Supplementary Table 1. Written informed consent was obtained from all individuals and the study protocol was approved by the Institutional Review Board of University General Hospital of Heraklion (#17652). The second cohort consisted of NSCLC individuals (Stages I and II) who fulfilled the criteria to undergo surgery for therapeutic purposes at the Thoracic Dept of Athens Chest Hospital “Sotiria” (*n* = 19). Individuals selected were treatment naïve at the time of surgery and heparinized peripheral blood and primary human lung tumor tissue were obtained. Of the 19 individuals with NSCLC, 14 were histopathologically annotated as lung adenocarcinoma, 4 as squamous cell carcinoma and 1 was not otherwise specified (NOS). Clinical characteristics including age, sex, smoking status, pathological subtypes and stages and comorbidities on all donors were recorded at recruitment and are listed in Table 2 and Supplementary Table 2. A third cohort of NSCLC individuals (Stages I-IV) underwent fibre-optic bronchoscopy with endobronchial biopsy (*n*=10). Clinical characteristics including age, sex, smoking status, pathological subtypes, and stages on all donors were recorded at recruitment and are listed in Table 3 and Supplementary Table 3. Regarding the cohort of individuals with bronchiectasis (Table 3), they also underwent fibre-optic bronchoscopy with endobronchial biopsy (*n*=10). The median age was 65.4, 60% were female and 50% current smokers. Prior to participation in the study, all individuals provided informed written consent and the protocol was approved by Athens Chest Hospital “Sotiria” research ethics committee [11504/20/04/21]. All studies were conducted according to the principles of the Declaration of Helsinki. The cohort that underwent thoracentesis was recruited through the Department of Critical Care and Pulmonary Medicine of Athens Evangelismos Hospital and consisted of individuals with NSCLC (*n*=3), small cell lung cancer (SCLC) (*n*=1), malignant mesothelioma (*n*=2) and pleural infection (*n*=4) (Supplementary Tables 4). Written informed consent was obtained from all individuals and the study protocol was approved by Evangelismos Hospital Ethics Committee (186/9-6-2022).

### PBMC and tumor infiltrating leukocyte cell fraction extraction

Peripheral blood mononuclear cells (PBMCs) were obtained from NSCLC individuals by density-gradient centrifugation (Lymphoprep, STEMCELL Technologies, Inc.). Primary lung tumor tissue was obtained from NSCLC individuals undergoing surgical resection. Tissues were minced and incubated with 0.14 units/ml Liberase TL (Sigma Aldrich) and 0.1mg/ml DNase I (Sigma Aldrich) in 37°C for 1h. Single-cell suspensions were prepared by passing the minced tissues through 70 µm cell strainers. Cells were resuspended in 40% percoll (Sigma Aldrich), laid over equal volume of 80% percoll and centrifuged at 600g for 25min without deceleration. The tumor infiltrating leukocyte cell fraction was collected at the interface between 40% and 80% discontinuous percoll gradient. PBMCs and the tumor infiltrating leukocyte cell fraction were cryopreserved in 90% FCS-10% DMSO freezing media until use.

#### Fibre-optic bronchoscopy and sample collection

Individuals with bronchiectasis (*n*=10) or with suspected NSCLC or known, previously treated, NSCLC, now suspected with possible relapse (*n*=10), underwent fibre-optic bronchoscopy with endobronchial biopsy EBUS TBNB and bronchoalveolar lavage fluid (BALF) collection. Bronchoscopy was performed on an outpatient basis, as previously described^53^. After inspection of the bronchial tree, bronchoalveolar lavage was performed and samples with a fluid recovery of ≥60% were retained for further analysis according to the ERS task force guidelines regarding measurements of acellular components in BAL^54^.

#### Pleural fluid collection

Individuals with NSCLC (*n*=3), malignant mesothelioma (*n*=2), SCLC (*n*=1) or pleural infection (*n*=4) underwent thoracentesis using a 21g needle for diagnostic or therapeutic purposes. Pleural fluid was collected in 4 ml Ethylenediaminetetraacetic Acid Tetrasodium-coated Vacutainer tubes. Fluid samples were centrifuged at 400 x g for 10 min and cell pellets were resuspended in 1ml of Bambanker (Nippon Genetics, alternatively in 10% DMSO in FBS) and stored at -80°C until analysis.

#### Mass cytometry – CyTOF

For high dimensional immunophenotyping analyses we utilised the Maxpar Direct Immunophenotyping Assay (MDIPA), which comprises of 30 pre-conjugated antibodies in lyophilized form, Standard Biotools, San Francisco, CA^55^. The MDIPA backbone was complemented with antibodies against CD33, PD-1, and CD274/PD-L1. The antibody panel used, clones and metal tags are provided in **Supplementary Table 5**. The antibodies against CD33, PD-1 and PD-L1 were titrated according to manufacturer’s instructions. For staining, PBMCs and TILs were thawed in prewarmed RPMI supplemented with 10% FBS, washed twice and then resuspended in fresh medium. For live/dead cell discrimination, cells were stained with 1 μM Cisplatin Cell-ID™ (Standard Biotools, San Francisco, CA) and washed with Maxpar cell staining buffer (CSB) followed by a blocking step (Human TruStain FcX, Biolegend). Then, cells were stained for cell surface markers with the MDIPA backbone as well as CD33, PD-1 and PD-L1 according to manufacturer’s instructions followed by two washes with CSB and fixation (1.6% filtered formaldehyde solution, Sigma) for 20 min at RT. Finally, cells were stained in DNA intercalator solution (1:1000 dilution of 125 μM Cell-ID™ Intercalator-Ir,) in Maxpar Fix and Perm buffer (all from Standard BioTools, San Francisco, CA). The following day, cells were washed with CSB buffer and Cell Acquisition Solution (CAS). Immediately before acquisition, cells were resuspended with EQ Passport beads (1:10 dilution). To maximize data quality, acquisition rate on the Helios™ system (Standard BioTools, South San Francisco, CA, USA) was maintained at a rate of 350 to 400 events/s. Acquired data were normalized using Passport beads (Standard BioTools method) with CyTOF software (version 10.7.1014). Prior to analysis, we performed data clean-up for gaussian parameters and live singlet cell events were used for downstream analyses. For these analyses, we used bivariate dot plots in FlowJo™ v10.8 Software (BD Life Sciences, Franklin Lakes, NJ, USA). FlowSOM clustering analysis and dimensionality reduction with UMAP were performed in R programming environment (Version 4.1.0, https://www.r-project.org/) following previously published, open source, validated workflows^25^. For each sample, data from living CD45^+^ cells were selected and imported into R using the flowCore package v2.4.0. All CyTOF data were transformed using hyperbolic *arcsinh* with a cofactor of 5. The initial clustering and dimensional reduction were performed using the FlowSOM and UMAP algorithms, respectively, with 33 lineage markers to accurately identify the major populations. Next, memory CD8^+^ T cells were selected by excluding the naïve cell cluster (T8 N; CD45RA^+^ CCR7^+^ positive cells) and reanalysed separately using a panel of markers including CD45RO, CD45RA, CCR7, PD-1, CXCR3, PD-L1, CCR4, CD28, CD27, CD16, CD57, CCR6, CD38, HLA-DR, CD294, CD161, and CD56.

#### Imaging Mass Cytometry (IMC) sample preparation and labelling

Formalin-fixed paraffin-embedded lung tissues sections were dewaxed in xylene for 20 min, hydrated in descending grades of ethanol (100%, 90%, 70%) and washed in distilled water for 5 min. Subsequently, tissues were incubated for 20 min at 98°C in preheated antigen retrieval solution (Dako Target Retrieval Solution, pH 9). Afterwards, tissues were let to cool down at room temperature for 10 min and then washed in distilled water for 10 min. Next, tissues were incubated with PBS with Calcium and Magnesium (PBS+/+) supplemented with 3% BSA, 0.1% Triton X-100, 5% normal mouse serum, 5% normal rabbit serum and 5% normal goat serum for 45 min at room temperature in a hydration chamber. To prepare the antibody cocktail, the total volume of antibodies at concentrations specific for the assay were calculated and diluted in PBS+/+ supplemented with 0.3% BSA, 0.01% Triton X-100, 0.5% normal mouse serum, 0.5% normal rabbit serum and 0.5% normal goat serum. Tissues were then incubated overnight with the antibody cocktail at 4 °C in a hydration chamber. All the carrier-free purified primary antibodies used for metal conjugation are shown in **Supplementary Table 5**. Following the overnight incubation, tissues were washed 4 times in 0.05% Tween-20 in PBS for 5 min and stained with Intercalator-Ir191/193, 0.3 mM, in PBS for 30 min at room temperature in a hydration chamber. Next, tissues were washed 3 times in 0.05% Tween-20 in PBS for 3 min. Finally, tissues were washed in Ultrapure water for 30 seconds and airdry for 20 min.

#### IMC data acquisition and analysis

Image acquisition was performed using the Hyperion imaging mass cytometry system (Standard Biotools). The system was tuned routinely to account for machine performance variability following manufacturer’s instruction. The ROIs of choice were ablated by setting the laser frequency and power to 200Hz and 2, respectively. Data were exported as MCD files and visualized using the MCD viewer (version 1.0, Standard Biotools). Steinbock was used to convert the original MCD file to TIFF format, perform cell segmentation using the pre-trained deep learning model DeepCell, and consolidate all object data (e.g., intensities, region properties) from all images into a single file^56,57^ . The ImcRtools (v. 1.8.0) and cytomapper (v. 1.10.1) packages were employed to read the single-cell data extracted using the Steinbock framework. Batch correction and integration of cells across samples were carried out using the Harmony algorithm^58^ . The bluster package was used to cluster cells using a shared nearest neighbor (SNN) graph with *k*=30. Cell-cell spatial interactions were computed using the *testInteractions* function, which calculates the average cell type/cell type interaction count and compares it against an empirical null distribution generated by permuting all cell labels^59^. Imaging Mass Cytometry experiments have been performed by the Multiscale Immuno-Imaging Unit, IRCCS Humanitas Research Hospital, Rozzano, Milan, Italy.

#### Flow cytometry and cell sorting

PBMCs were stained for 30 minutes at 4°C with the following monoclonal anti-human antibodies: CD3-APC-Cy7 (OKT3), CD4-PerCP-Cy5.5 (OKT4), CD8-BV785 (RPA-T8), CD25-BV605 (BC96), CD127-BV510 (AO19D5), CD14-FITC (M5E2), CD16-PE (3G8), CD279 (PD1)-PE-Cy7 (A17188B), PD-L1-BV421 (29E.2A.3), HLA-DR-Alexa Fluor 700 (LN3), CD123-APC (6H6) all from Biolegend. BAL cells were stained for 30 minutes at 4°C with the following monoclonal anti-human antibodies: CD3-PE (HIT3a), CD8-PE-Cy7 (HIT8a), PD-1-FITC (EH12.2H7) and PD-L1-BV421 (29E.2A3) all from Biolegend. Flow cytometry acquisition and analysis was performed with a BD FACSAria III (BD Biosciences) and BD FACSCelesta. FACS sorting under aseptic conditions was performed using 70-micron nozzle and 4-way purity DIVA software sorting precision protocol. During cell sorting PD-L1 positive CD8 T cells were collected in 1.5 ml collection tubes following the sorting strategy as shown in **Supplementary** Figure 2. Analysis of flow cytometry data were done with FlowJo™ v10.8 Software (BD Life Sciences, Franklin Lakes, NJ, USA).

#### RNA Isolation and 3′ RNA sequencing

RNA from FACS-sorted CD8 PD-L1 positive cells (21×10^3^ on average) from responders (*n*=5) and non-responders (*n*=5) NSCLC individuals was isolated using Nucleospin RNA XS kit (Macherey-Nagel), according to the manufacturer instructions. The quality and quantity of isolated RNA was measured using 2100 Bioanalyser (Agilent) and RNA 6000 Pico Kit reagents (Agilent). RNA samples with RNA integrity number (RIN)>7 were used for library construction using the 3′ mRNA-Seq Library Prep Kit FWD for Illumina (QuantSeq-LEXOGEN) as per the manufacturer’s instructions. Amplification was controlled for obtaining optimal unbiased libraries across samples by assessing the number of cycles (ranging from 20-24) required by qPCR. DNA High Sensitivity Kit for bioanalyzer was used to assess the quantity and quality of libraries, according to the manufacturer’s instructions (Agilent). Libraries were multiplexed and sequenced on an Illumina Nextseq 500 at the genomics facility of IMBB FORTH according to the manufacturer’s instructions.

### Differential Expression Analysis (DEA) of bulk RNA sequencing data

The quality of the FASTQ files was assessed with the FastQC software. Reads were aligned to the human (hg38) genome with the Hisat2 aligner37 (hisat2 -p32 -x $REFERENCE_GENOME -q fastq/$FILE_ID.fastq -S $FILE_ID.sam --score-min L,0,-0.5 -k 2). Htseq-counts was utilized to summarize reads at the gene level (htseq-count -f bam -s yes -i gene_id bam/$FILE_ID.bam data/refs/Homo_sapiens/UCSC/hg38/Annotation/Genes/genes.gtf>$COUNTS_DIR/NGS$FILE_ID). Differential expression analysis (DEA) was conducted by running EdgeR^60^ via SARTools 1.5.0^61^. For the comparison of PD-L1+ vs. PD-L1− CD8 positive sorted cells, DEGs in either group of the comparison were defined by applying the following thresholds: |Log2FC| >1.5 and p-value <0.05, which was considered statistically significant. For the comparison of NR and R, PD-L1+ CD8 T cell samples were divided according to the RECIST 1.1 criteria. DEGs in either group were identified using the thresholds |Log2FC| > 0.5 and p-value < 0.05, which were considered statistically significant. Heatmaps and boxplots were created in R with an in-house–developed script which is based on the complex heatmap R package^62^.

### Motif enrichment analysis

The PScan software tool (version 1.6) was used for the in silico computational analysis of overrepresented TF binding sites within the 5′-promoter regions of genes upregulated within the PD-L1+ CD8+ T cells^63^ . PScan was executed on the [–950, +50] bp upstream regions using the Homo sapiens JASPAR 2018_NR database^64^ . The results were summarized with a scatter plot, where p-values were plotted against Z-scores on the vertical axis using GraphPad Prism 8.

### Gene set enrichment analysis (GSEA) and motif enrichment analysis

GSEA was performed in R using the fgsea package (v. 1.24.0) with a gene list ranked by log2 fold changes. The analysis was conducted in preranked mode with 1,000 permutations. The maximum gene set size was set to 500 genes, and the minimum gene set size was set to 15 genes. The gene signature was retrieved from the H collection (h.all.v7.5.1.symbols.gmt) of the Molecular Signatures Database (MSigDB v7.5.1)^65^.

### Survival analysis

DEGs identified between R and NR samples were used to generate survival curves. The Kaplan-Meier Plotter online tool^66^ was employed for statistical analysis and survival curve generation, utilizing the NSCLC immunotherapy cohort.

### PENCIL and Stabl prediction analysis

The PENCIL method is based on LWR, a machine learning technique that incorporates rejection labels into prediction results^26^. For PENCIL, we used hyperbolic arcsine-transformed single-cell mass cytometry intensity matrices along with relevant cell metadata, such as cluster ID and UMAP coordinates of T8 memory cells. These data were sourced from CATALYST and imported into Seurat^67^ (v. 5.0.3) via the *CreateSeuratObject* function. We adhered to the standard workflows for making predictions as outlined on the PENCIL GitHub page. Analyses with PENCIL were performed using Python 3.11.4 with GPU acceleration (NVIDIA Quadro RTX 5000), and default settings were applied for the shuffle rate, lambda L1, and lambda L2 tuning parameters. For StablL feature selection, we used the binary classification mode (Jupyter Notebook) with the matrix containing the T8 memory cluster frequencies as input values. The following tuning parameters were used for the analysis: lambda_grid = “0.1, 1, 30”; hard_threshold = 0.5; random_state= 42; n_bootstraps=1,000; fdr_threshold_range= “0.1, 1, 0.01”.

### In silico scRNA-seq data analysis

Tumor infiltrating CD8+ T cells were retrieved from GSE99254(NSCLC, stage I-IV), GSE120575 (NSCLC, stage I-IV), GSE154826 (NSCLC, stage I-IIB) dataset. All three datasets have been normalized using the R package Seurat (v. 5.0.3) applying SCTransform function^68^. For GSE99254 and GSE120575 dataset CD8+ T cells were annotated as PD-L1^+^ applying the following thresholds on the expression level of CD274 gene (coding for PD-L1) > 3. Cells with CD274 below of threshold (<3) were classified as PD-L1^−^. Since the expression of CD274 was not retrieved for the dataset GSE154826 due to low sequencing depth, we proceeded with the standard workflow by analyzing both the transcripts and cell-surface proteins present within this dataset (CITE-seq technology). Surface protein expression was normalized using the ’CLR’ method. We computed the principal components (PCs) and selected 20 PCs to run UMAP and perform graph-based clustering with a resolution ranging from 0.1 to 1. Clustertree was used to determine the optimal resolution^69^. Differences in gene expression between PD-L1^+^ and PD-L1^−^ CD8^+^ T cells were assessed using a Wilcoxon Rank Sum test.

## Supplementary Figure legends

**Supplementary Fig. 1.**
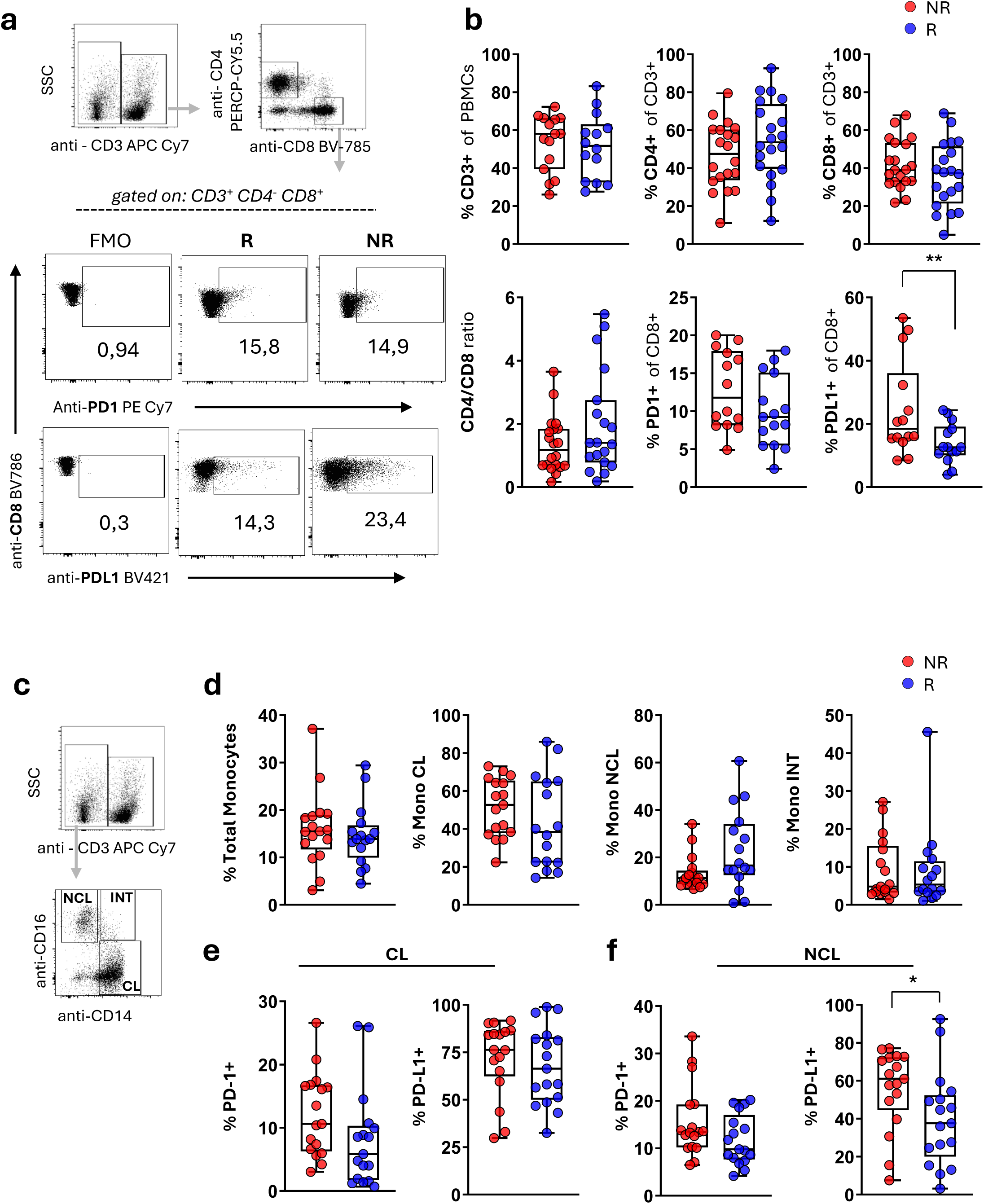
| Flow cytometry analysis reveals increased percentages of PD-L1^+^ CD8^+^ T cells in the peripheral blood of NSCLC patients resistant to anti-PD1 immunotherapy. **a)** Dot plots showing gating strategy to analyse CD4^+^ and CD8^+^ T cells in PBMCs from NSCLC patients before the initiation of aPD1 immunotherapy, responders (R) and non-responders (NR) to aPD1 immunotherapy and gating strategy for PD1 positive and PD-L1 positive CD8 T cells. (FMO : Fluorescence minus one control) **b)** Box and whiskers plots of collective flow cytometry data of PBMC from NSCLC patients (n=20 patients per group), showing the abundance of total CD3 T cells, the percentage of CD4 and CD8 subpopulations, the CD4/CD8 ratio and the percentage of PD1, PD-L1 CD8^+^ T cells. **c)** Representative FACS Dot plots for each group of individuals showing gating strategy for CD8+ PD-L1+ T cells. Unpaired two tailed Mann-Whitney test **p <0.01.

**Supplementary Fig. 2.**
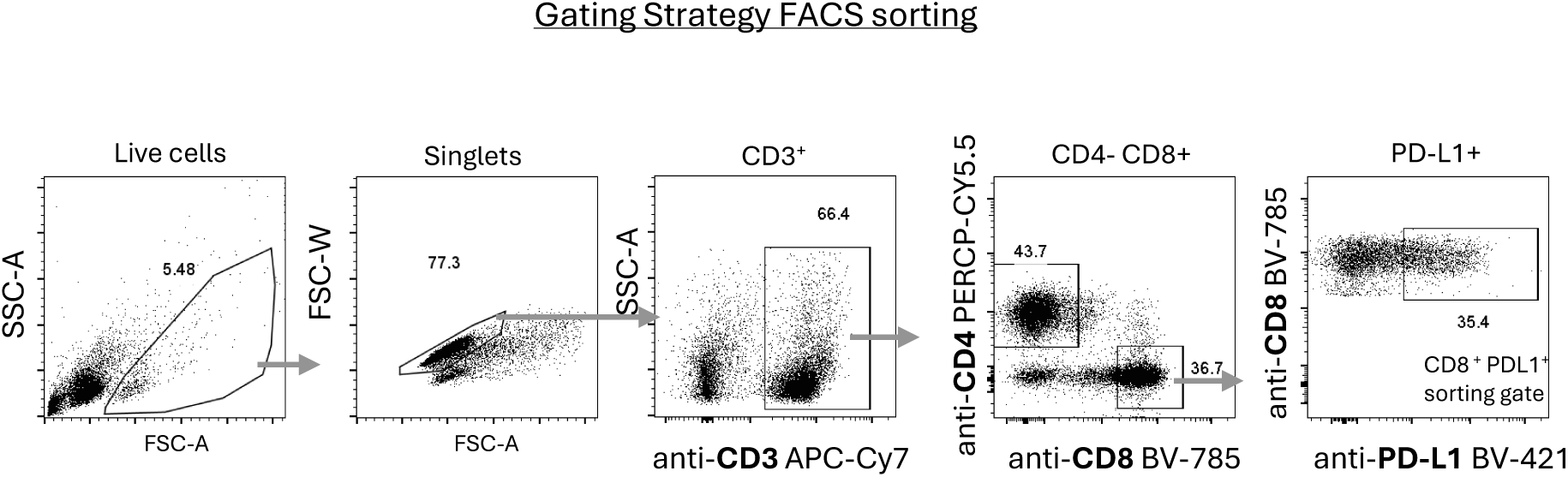
| Gating strategy for flow cytometric sorting of CD8+ PD-L1+ cells from peripheral blood of NSCLC patients. Representative FACS Dot plots of the gating strategy to sort live (SSC-A *vs* FSC-A plot), singlets (FSC-W *vs* FSC-A plot), CD3^+^ (SSC-A *vs* CD3 plot), CD8^+^ (CD4 *vs* CD8 plot), PD-L1^+^ cells (CD8 *vs* PD-L1 plot) from PBMCs of NSCLC patients.

## Data Availability

All data produced in the present study are available upon reasonable request to the authors.

## AUTHOR CONTRIBUTIONS

Conceptualized the study: PV Designed the study: MS, NP and PV

Performed experiments and analyzed data: NP, DLT, AT, PS, AM, EM, AV, IM, SM

Provided clinical samples and patients information: CF, KD, IK, KS, KP, DM

Wrote and edited the manuscript: MS, NP, DLT, AC, SDB and PV

## Acknowledgements

We acknowledge support of this work by the project “The Greek Research Infrastructure for Personalised Medicine (pMedGR)” (MIS 5002802) to P.V., which is implemented under the Action “Reinforcement of the Research and Innovation Infrastructure”, funded by the Operational Programme “Competitiveness, Entrepreneurship and Innovation” (NSRF 2014-2020) and co-financed by Greece and the European Union (European Regional Development Fund).

This work was further supported by the “Infrastructure for Preclinical and early-Phase Clinical Development of Drugs, Therapeutics and Biomedical Devices (EATRIS-GR)” (MIS 5028091), which is implemented under the Action “Reinforcement of the Research and Innovation Infrastructure”, funded by the Operational Programme “Competitiveness, Entrepreneurship and Innovation” (NSRF 2014-2020) and co-financed by Greece and the European Union (European Regional Development Fund). DLT is research fellow supported by the Associazione Italiana per la Ricerca sul Cancro (AIRC IG ID 25073, to AC). SDB is supported by Associazione Italiana per la Ricerca sul Cancro (MFAG 2023 ID 29036). This work was also supported from the Hellenic Foundation for Research and Innovation (H.F.R.I.) under the action “Always stive for Excellence-Theodoros Papazoglou” grant # 1429 to P.V.

