## Supplementary Table 5 for "Blood immunomap for prediction of responses to aPD1 immunotherapy in metastatic non-small cell lung cancer"

**Supplementary Table X.** **Antibodies used for CyTOF**

|  | **Target** | **Clone** | | **Metal** | **Source** | **Localization** | **Dilution** |
| --- | --- | --- | --- | --- | --- | --- | --- |
| 1 | **CD45** | HI30 | | 89 Y | MDIPA | Cell-surface | - |
| 2 | **CCR6** | G034E3 | | 141 Pr | MDIPA | Cell-surface | - |
| 3 | **CD123** | 6H6 | | 143 Nd | MDIPA | Cell-surface | - |
| 4 | **CD19** | HIB19 | | 144 Nd | MDIPA | Cell-surface | - |
| 5 | **CD4** | RPA‐T4 | | 145 Nd | MDIPA | Cell-surface | - |
| 6 | **CD8a** | RPA‐T8 | | 146 Nd | MDIPA | Cell-surface | - |
| 7 | **CD11c** | Bu15 | | 147 Sm | MDIPA | Cell-surface | - |
| 8 | **CD16** | 3G8 | | 148 Nd | MDIPA | Cell-surface | - |
| 9 | **CD45RO** | UCHL1 | | 149 Sm | MDIPA | Cell-surface | - |
| 10 | **CD45RA** | HI100 | | 150 Nd | MDIPA | Cell-surface | - |
| 11 | **CD161** | HP‐3G10 | | 151 Eu | MDIPA | Cell-surface | - |
| 12 | **CCR4** | L291H4 | | 152 Sm | MDIPA | Cell-surface | - |
| 13 | **CD25** | BC96 | | 153 Eu | MDIPA | Cell-surface | - |
| 14 | **CD27** | O323 | | 154 Sm | MDIPA | Cell-surface | - |
| 15 | **CD57** | HCD57 | | 155 Gd | MDIPA | Cell-surface | - |
| 16 | **CXCR3** | G025H7 | | 156 Gd | MDIPA | Cell-surface | - |
| 17 | **CXCR5** | J252D4 | | 158 Gd | MDIPA | Cell-surface | - |
| 18 | **CD28** | CD28.2 | | 160 Gd | MDIPA | Cell-surface | - |
| 19 | **CD38** | HB‐7 | | 161 Dy | MDIPA | Cell-surface | - |
| 20 | **CD56** | NCAM16.2 | | 163 Dy | MDIPA | Cell-surface | - |
| 21 | **TCRgd** | B1 | | 164 Dy | MDIPA | Cell-surface | - |
| 22 | **CD294** | BM16 | | 166 Er | MDIPA | Cell-surface | - |
| 23 | **CCR7** | G043H7 | | 167 Er | MDIPA | Cell-surface | - |
| 24 | **CD14** | 63D3 | | 168 Er | MDIPA | Cell-surface | - |
| 25 | **CD3** | UCHT1 | | 170 Er | MDIPA | Cell-surface | - |
| 26 | **CD20** | 2H7 | | 171 Yb | MDIPA | Cell-surface | - |
| 27 | **CD66b** | G10F5 | | 172 Yb | MDIPA | Cell-surface | - |
| 28 | **HLADR** | | LN3 | 173 Yb | MDIPA | Cell-surface | - |
| 29 | **IgD** | | IA6‐2 | 174 Yb | MDIPA | Cell-surface | - |
| 30 | **CD127** | | A019D5 | 176 Yb | MDIPA | Cell-surface | - |
| 31 | **CD33** | | WM53 | 169 Tm | SB | Cell-surface | 0.4/300  (stock: 0.5 mg/ml) |
| 32 | **CD274/**  **PD-L1** | | MIH1 | 209 Bi | SB | Cell-surface | 1/300  (stock: 0.5 mg/ml) |
| 33 | **CD279/**  **PD-1** | | EH12.2H7 | 165 Ηο | SB | Cell-surface | 1/300  (stock: 0.5 mg/ml) |
| 34 | **a-SMA** | | 1A4 | 141Pr | SB | Intracellular | - |
| 35 | **Vimentin** | | D21H3 | 143Nd | SB | Intracellular |  |
| 36 | **CD14** | | EPR3653 | 144Nd | SB | Cell-surface |  |
| 37 | **CD16** | | EPR16784 | 146Nd | SB | Cell-surface |  |
| 38 | **Pan-Keratin** | | C11 | 148Nd | SB | Intracellular |  |
| 39 | **PD-L1** | | E1L3N | 150Nd | SB | Cell-surface |  |
| 40 | **CD57** | | NK/804 | 152Sm | SB | Cell-surface |  |
| 41 | **CD11c** | | EP1347Y | 154Sm | SB | Cell-surface |  |
| 42 | **FoxP3** | | PCH101 | 155Gd | SB | Intracellular |  |
| 43 | **CD4** | | EPR6855 | 156Gd | SB | Cell-surface |  |
| 44 | **E-Cadherin** | | 24E10 | 158Gd | SB | Cell-surface |  |
| 45 | **CD68** | | KP1 | 159Tb | SB | Intracellular |  |
| 46 | **CD20** | | H1 | 161Dy | SB | Cell-surface |  |
| 47 | **CD8a** | | D8A8Y | 165Ho | SB | Cell-surface |  |
| 49 | **PD-1** | | EPR4877(2) | 156Gd | SB | Cell-surface |  |
| 50 | **granzyme B** | | DO-7 | 143Nd | SB | Intracellular |  |
| 43 | **Ki-67** | | B56 | 168Er | SB | Intracellular |  |
| 44 | **Collagen I** | | polyclonal | 169Tm | SB | Cell-surface |  |
| 45 | **CD3** | | polyclonal | 170Er | SB | Cell-surface |  |
| 46 | **CD45RO** | | UCHL1 | 173Yb | SB | Cell-surface |  |
| 47 | **HLA-DR** | | LN3 | 174Yb | SB | Cell-surface |  |
| 49 | **Histone H3** | | D1h2) | 176Yb | SB | Intracellular |  |

Abbreviations: MDIPA; Maxpar Direct Immune Profiling Assay from SB, SB; Standard Biotools Inc. (formerly Fluidigm)
