## Supplementary material for "Blood immunomap for prediction of responses to aPD1 immunotherapy in metastatic non-small cell lung cancer": Table 1

**Table 1**. NSCLC (stage III-IV) patients characteristics.

| Characteristic (N=56) N (%) | |
| --- | --- |
| Age |  |
| Median | 68 |
| Range | 37-87 |
| Gender |  |
| Female | 10 (17.9) |
| Male | 46 (82.1) |
| Histology |  |
| Adenocarcinoma | 36 (64.3) |
| Squamous cell carcinoma | 17 (30.4) |
| Pleomorphic carcinoma | 2 (3.5) |
| Adenocarcinoma & Squamous cell carcinoma | 1 (1.8) |
| Smoking status |  |
| Current | 23 (41.1) |
| Former | 22 (39.3) |
| Never | 2 (3.5) |
| N/A | 9 (16.1) |
| TNM Stage |  |
| IIB | 1 (1.8) |
| IIIA | 1 (1.8) |
| IIIB | 7 (12.5) |
| IIIC | 1 (1.8) |
| IV | 46 (82.1) |
| Treatment |  |
| Nivolumab | 10 (17.9) |
| Durvalumab | 5 (8.9) |
| Pembrolizumab | 3 (5.36) |
| Pembrolizumab/Alimta/Carboplatin | 18 (32.1) |
| Pembrolizumab/Abraxane/Carboplatin | 10 (17.9) |
| Pembrolizumab/Paclitaxel/Carboplatin | 3 (5.36) |
| Pembrolizumab/Alimta/CDDP | 2 (3.5) |
| Atezolizumab/Abraxane/Carboplatin | 1 (1.8) |
| Ipilimumab/Nivolumab/Alimta/Carboplatin | 3 (5.36) |
| Ipilimumab/Nivolumab/ Paclitaxel/Carboplatin | 1 (1.8) |
| Response (RECIST 1.1) |  |
| Responders | 29 (51.8) |
| Non-Responders | 27 (48.2) |
