## Supplementary material for "Blood immunomap for prediction of responses to aPD1 immunotherapy in metastatic non-small cell lung cancer": Table 2

**Table 2**. NSCLC (stage I-II) patients characteristics.

| Characteristic (N=19) N (%) | |
| --- | --- |
| Age |  |
| Median | 69 |
| Range | 50-82 |
| Gender |  |
| Female | 9 (47.4) |
| Male | 10 (52.6) |
| Histology |  |
| Adenocarcinoma | 14 (73.7) |
| Squamous cell carcinoma | 4 (21.0) |
| NOS NSCLC | 1 (5.3) |
| Smoking status |  |
| Current | 11 (57.9) |
| Former | 6 (31.6) |
| Never | 2 (10.5) |
| TNM Stage |  |
| IA | 5 (26.3) |
| IB | 3 (15.8) |
| IIA | 4 (21.1) |
| IIB | 7 (36.8) |
| Comorbidities |  |
| Heart Disease | 9 (47.4) |
| Diabetes | 4 (21.0) |
| Chronic Obstructive Pulmonary Disease | 1 (5.3) |
| Autoimmune disorders | 1 (5.3) |
| Allergic disorders | 1 (5.3) |
| Depression | 2 (10.5) |
| N/A | 1 (5.3) |
