## Supplementary material for "Blood immunomap for prediction of responses to aPD1 immunotherapy in metastatic non-small cell lung cancer": Table 3

**Table 3**. NSCLC (stage I-IV) patients characteristics.

| Characteristic (N=10) N (%) | |
| --- | --- |
| Age |  |
| Median | 74 |
| Range | 52-87 |
| Gender |  |
| Female | 4 (40) |
| Male | 6 (60) |
| Histology |  |
| Adenocarcinoma | 6 (60) |
| Squamous cell carcinoma | 3 (30) |
| Pulmonary sarcomatoid carcinoma | 1 (10) |
| Smoking status |  |
| Current | 9 (90) |
| Former | 1 (10) |
| Never | 0 (0) |
| TNM Stage |  |
| IA | 2 (20) |
| IIIB | 1 (10) |
| IV | 8 (80) |
